# Virtual Trials: Causally-validated treatment effects efficiently learned from an observational cancer registry

**DOI:** 10.1101/2021.06.12.21258409

**Authors:** Asher Wasserman, Al Musella, Mark Shapiro, Jeff Shrager

## Abstract

Randomized controlled trials (RCTs) offer a clear causal interpretation of treatment effects, but are inefficient in terms of information gain per patient. Moreover, because they are intended to test cohort-level effects, RCTs rarely provide information to support precision medicine, which strives to choose the best treatment for an individual patient. If causal information could be efficiently extracted from widely available real-world data, the rapidity of treatment validation could be increased, and its costs reduced. Moreover, inferences could be made across larger, more diverse patient populations. We created a “virtual trial” by fitting a multilevel Bayesian survival model to treatment and outcome records self-reported by 451 brain cancer patients. The model recovers group-level treatment effects comparable to RCTs representing over 3200 patients. The model additionally discovers the feature-treatment interactions needed to make individual-level predictions for precision medicine. By learning from heterogeneous real-world data, virtual trials can generate more causal estimates with fewer patients than RCTs, and they can do so without artificially limiting the patient population. This demonstrates the value of virtual trials as a complement to large randomized controlled trials, especially in highly heterogeneous or rare diseases.

## Randomized Controlled Trials are inefficient

Cancer is a complex family of diseases, with a high degree of variation in outcomes. The challenge of precision oncology is to predict the effect of treatments on outcomes, conditional on features unique to a given patient. Randomized controlled trials (RCTs) are often considered the “gold standard” method for inferring the causal effect of medical interventions. However, an RCT can only test a handful of hypotheses. Relevant patient features (hereafter generically referred to as biomarkers) potentially number in the hundreds or thousands, including clinical demographic features, and cancer stage, grade, morphology, histopathology, and genetics. Genetics has led to an explosion of possible biomarkers that may be relevant for cancer diagnosis, prognosis, or treatment. OncoKB, for example, lists 47 prognostic, 132 diagnostic, and 119 therapeutically relevant genes for cancer (*1*). The space of potential treatments is also vast, with hundreds of plausible treatments, and thousands of possible combinations. The combined space of biomarkers, treatments, and the pairwise interactions between these easily numbers in the thousands of dimensions, and the dimensionality increases rapidly as new biomarkers and treatments are discovered (*2, 3*). Indeed, the relevance of a trial is often reduced during the time required to conduct it, as new information emerges.

The availability of patients to populate RCTs is also a significant limitation on their efficiency. As the targeting of therapies becomes more narrowly focused, the number of patients matching the inclusion criteria becomes smaller, and factors such as inaccessibility, ineligibility, patient reluctance, and racial bias end up turning most patients away from clinical trials (*4*). In oncology, this is compounded by the relative over-abundance of trials in common cancers, and paucity of trials for rare and pediatric cancers. The net effect is that overall clinical trial participation in cancer is just 8.1% (*5*), and this figure is lower for minorities, children and the elderly. Thus, more than 90% of the potential data from which clinically-actionable information might be learned is not accessed by RCTs.

Finally, it is estimated that as many as 75% of all drugs are prescribed off-label, while for rare diseases and cancer, off-label usage can reach 90% (*6*). RCTs often measure a subpopulation which differs significantly from the ultimate recipients of the treatments.

### Modeling real world survival data

With the emergence of electronic health records and increased adoption of real world data, it is increasingly possible to collect and analyze data on a much larger and more diverse population. At the same time, advances in computing power and analytic tools provide new approaches for causal inference in large clinical datasets.

Precision medicine requires inferential tools that support informed individual treatment decisions. These tools must allow for continuous statistical learning from new data at the individual level, and from external sources of information. These inference methods must accommodate larger and more heterogeneous datasets. Because of the high-dimensional nature of this data, feature selection techniques are required to identify biomarkers that may be prognostically or therapeutically relevant, and the inferential process must produce results that are intuitively interpretable and can be understood by clinicians and explained to patients.

Biomarkers may affect disease prognosis, treatment response, or both. While the Cox proportional hazards (Cox PH) model has been the most widely used for survival analyses in cancer, deviations from the proportional hazards assumption are now more frequently observed in precision oncology trials (*7*). Further, the “hazard function” reported in the Cox PH model is ill-suited for patient-level prediction (*8*) because the proportionality assumption is violated when adding mediator variables which can shift baseline hazards rather than just altering the slope of the hazard function (*9*). Other issues with the basic Cox PH model are its inability to use time-dependent covariates as well as the chance for collinearity with high dimensional data, although recent progress has been made in addressing these issues (*10–12*).

In contrast, the accelerated failure time (AFT) model evaluates (log) survival time directly as a linear combination of covariates with an error term, making this model better suited to individual patient-level survival prediction models. Use of parametric AFT models has been explored in cancer (*13, 14*) and these models are able to accommodate larger numbers of covariates that affect individual predictions. AFT models may also be less sensitive to omitted covariates, more accommodating of time-dependent acceleration factors, and more amenable to causal inference (*15, 16*).

To have clinical utility, machine learning models that predict individual outcomes from a large feature and treatment-by-feature interaction space must be easily interpretable by a clinician without specialized statistical expertise. Because AFT models directly measure the effect of an explanatory variable on (log) survival time, rather than a hazard ratio, the measured effects of covariates has a more natural interpretation than survival models based on Cox PH, in which explanations of changes in hazard ratio can be misleading and confusing to patients (*17, 18*). Finally, when interpreting models, the Bayesian posterior probability distribution has a more intuitive interpretation than a confidence interval or p-value, which are based on the probability that hypothetical, counterfactual data from the null hypothesis would be more extreme than the observed data (*19*).

In this work, we present a Bayesian multilevel AFT model of patient outcomes with sparsity-inducing priors that may be used to learn from historical patient outcomes data to make predictions for the outcomes of new patients under proposed treatments. We apply this model to an observational dataset of patient-reported outcomes from patients with primary brain tumors. This model makes easy-to-interpret estimates of biomarker effects that contribute to predicted outcomes. We compare the degree of causal bias in the resulting model by comparing predictions of the model to data reported from several large RCTs.

### A Bayesian precision oncology model

Here we present our time-to-event model for learning treatment effects in observational data. The event in question may be any well-defined point in the time course of a disease. In the context of cancer, common endpoints include overall survival (OS; the time from treatment to death) and progression-free survival (PFS; the time from treatment to disease progression or death). Disease progression is often indicated by a growth in the tumor (or some proxy measurement) by some predefined factor.

A natural concern for any observational data model is the robustness of inference in the presence of unmeasured covariates. To mitigate this concern, we adopt a multilevel accelerated failure time (AFT) model (*15*). There are two levels in the model: one for patients, and one for treatment time interval. Each patient can be associated with one or more treatment time intervals. In AFT survival models, the effect of a treatment is to stretch or shrink the expected survival by a constant factor. Such a model may be viewed as a generalized linear model in which the outcome variable is the log of the survival time.

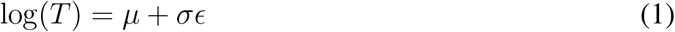

In equation 1, exp (*µ*) is the scale factor for the survival distribution, *σ* represents the variation in survival times about the expected survival time, and *ϵ* is a standardized distribution of error terms. Some common parametric choices for the distribution of *ϵ* are Logistic, Gumbel, and Normal, corresponding to Log-Logistic, Weibull, and Log-Normal distributions for the survival times, however a non-parametric spline-based approach may also be appropriate (*16*). Here we chose a Logistic distribution for *ϵ*, with standardized probability distribution function (PDF)

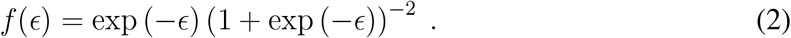

We assessed the appropriateness of the Log-Logistic distribution using non-parametric survival estimates and visual inspection of the relationship between survival time and survival probability (*20*).

To model the effect of patient features and treatments on observed outcomes, we expressed the scale and variance parameters of equation 1 as linear responses of patient-level predictors (subscript 1) and treatment-level predictors (subscript 2). The parameters of the survival time distribution of patient *i* during treatment time interval *j* are

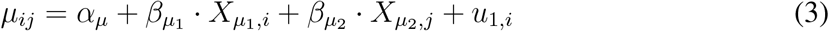

and

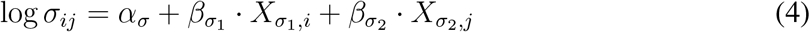

where *α*_{*µ,σ*}_ is the scalar intercept, 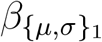 is the vector of effect sizes for patient-level predictors, 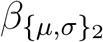 is the vector of effect sizes for treatment-level predictors, 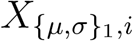 is the vector of predictors for the *i*-th patient, and 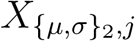 is the vector of predictors for the *j*-th treatment time interval. The term *u*_1,*i*_ refers to the patient-level random effect size parameter for the *i*-th patient, and they are assumed to be drawn from a Normal distribution with a mean of zero and a standard deviation of *σ*_*u*_. A graphical summary of the model using plate notation is shown in Figure 1.

**Figure 1:**
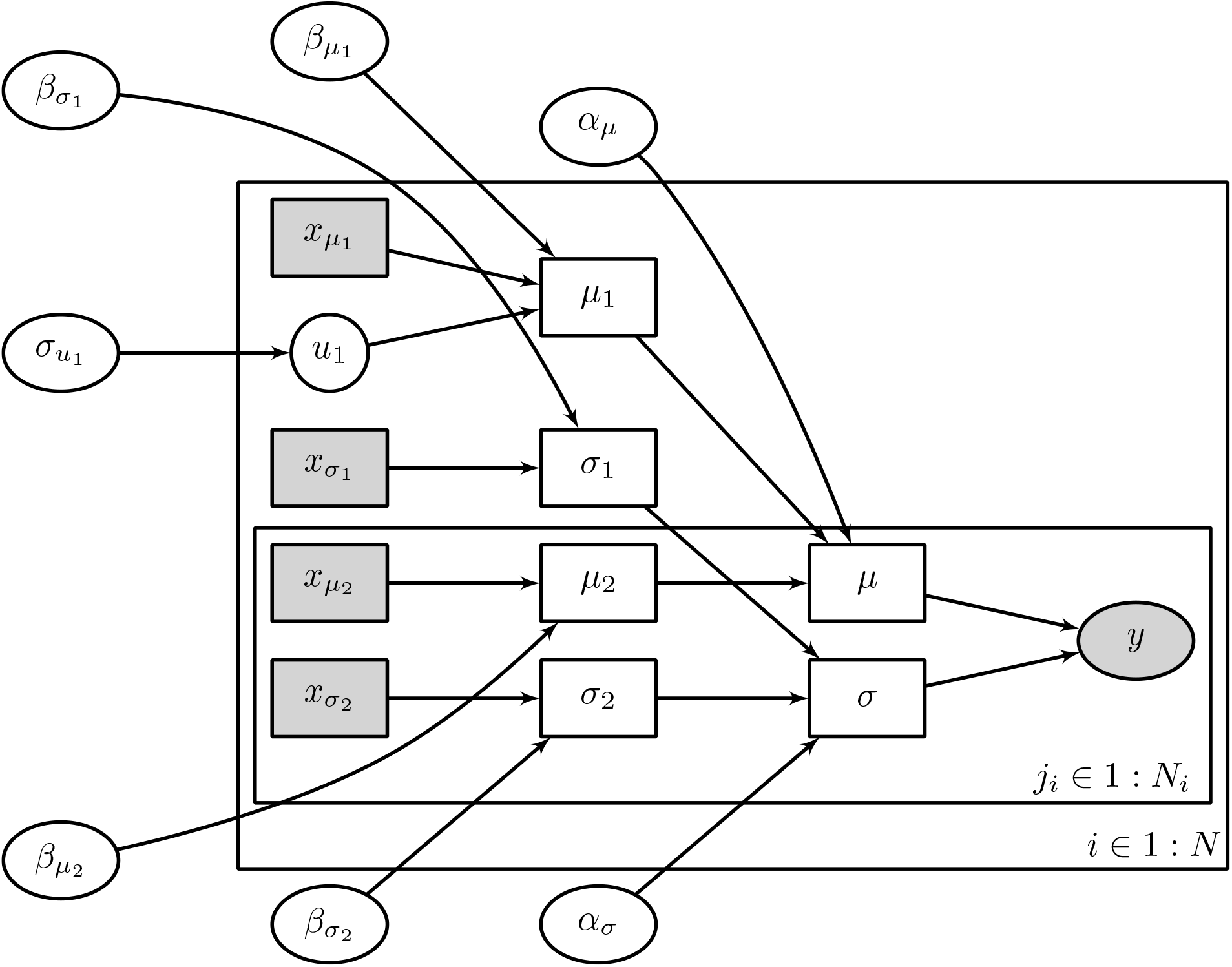
Graphical model representation using plate notation for the Bayesian multilevel model. Open ellipse nodes show latent parameters, filled ellipse nodes show observed outcome data, filled boxes show observed covariates, and open boxes show deterministically computed quantities. The outer plate (indexed by *i*) shows patient-level variables, including predictors for the expected survival time 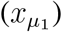, predictors for the variance 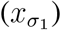, and patient-level random effects (*u*_1_). The inner plate (indexed by *j*_*i*_) shows treatment-level variables, including predictors for expected survival 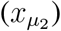 and predictors for the variance 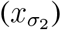.

Equation 1, together with a choice of probability distribution, *f* (*ϵ*), defines a likelihood for a set of survival times. For right-censored survival times, the likelihood is modified such that

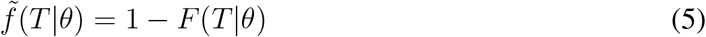

where *F* (*T* |*θ*) is the cumulative distribution of survival times, conditional on model parameters *θ*.

To regularize the regression fitting process, we defined prior distributions over all of the model parameters. For the intercept parameters *α* (as well as slopes for the log variance pa-rameters, *β*_*σ*_), we used a zero-centered Normal prior distribution with standard deviation *s*, a hyperparameter chosen to be some large but reasonably finite value. Following conventional practice in Bayesian multilevel modeling (*21, 22*), we used a Normal distribution over patient-level effects, *u*, whose scale *σ*_*u*_ is drawn from a standard Half-Cauchy distribution. For the slope parameters *β*_*µ*_, we anticipated a set of sparse effects; i.e., of the large number of predictors and interactions between predictors, we assumed that many of the associated effects would be below the noise threshold, but some fraction will have significant effects on the survival time. Thus, we used a regularized horseshoe prior (*23*) to induce sparsity in the joint distribution of effect size parameters, *β*_*µ*_. This sparsity-inducing prior allows us to model the full range of pairwise interaction terms and their effects on survival, without making specific assumptions about which terms to keep and which to discard.

### Causal interpretation

Figure 2 shows a graphical causal model interpretation of the model. As in the previous section, *x*_1_ refers to measured patient-level pre-treatment variables, while *x*_2_ refers to treatment-level variables. Unmeasured patient-level variables are denoted by *u*_1_. Each of *x*_1_, *x*_2_, and *u*_1_ can affect outcomes *y*, and *x*_1_ and *u*_1_ can affect *x*_2_. Thus *x*_1_ and *u* represent confounders for the effect of *x*_2_ on *y*. We would like to estimate the effect of *x*_2_ on *y* in an interventional scenario, where we force *x*_2_ to some value independent of *x*_1_ and *u*_1_ (visualized in Figure 2 as the removal of the red arrows).

**Figure 2:**
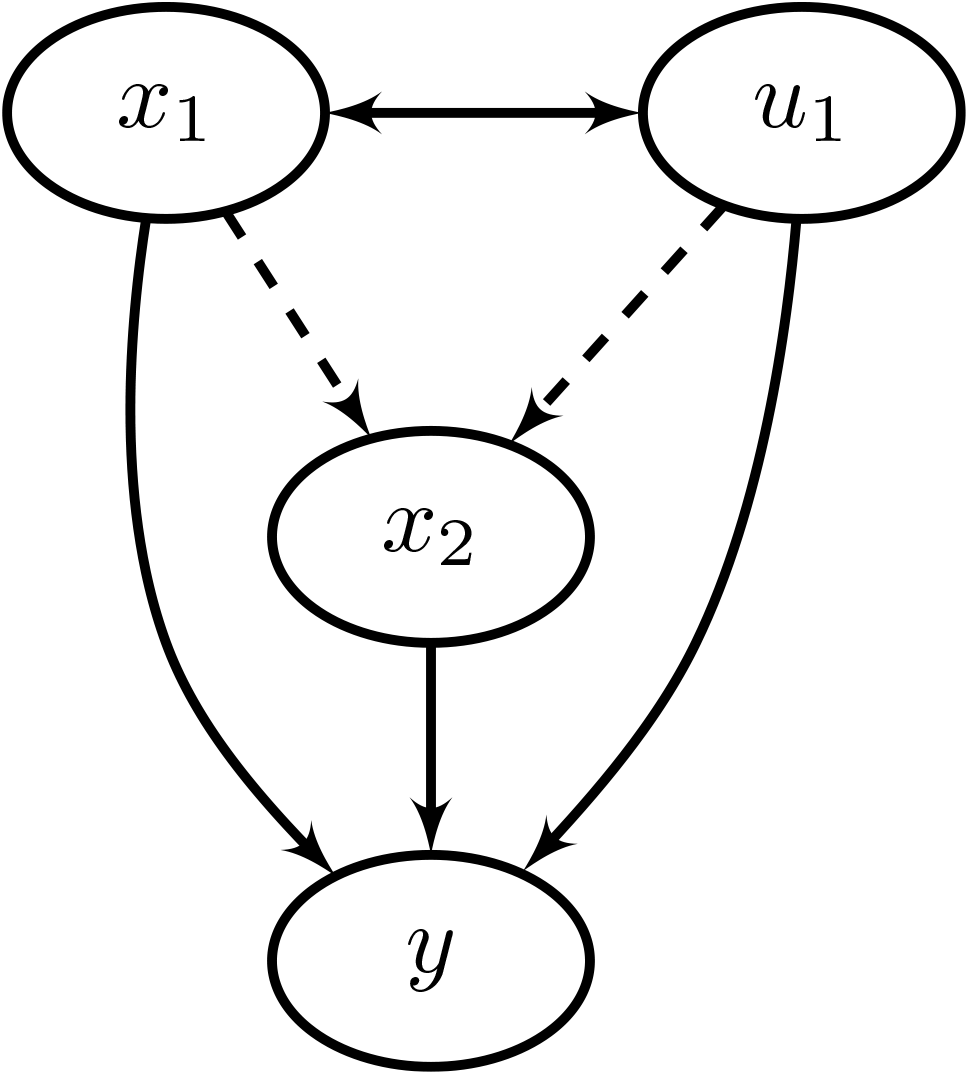
Graphical causal model relating measured pre-treatment variables (*x*_1_), unmeasured pre-treatment variables (*u*_1_), treatment variables (*x*_2_), and treatment outcomes (*y*). *x*_1_ and *u*_1_ are assumed to be unaffected by the choice of treatment, but they may have an effect on both the choice of treatment and the treatment outcome, and thus represent confounders on the effect of *x*_2_ on *y*. Removing the dashed arrows leading into *x*_2_ would yield the interventional causal graph associated to forcing *x*_2_ to some particular value.

Pearl’s backdoor criterion (*24*) specifies when we can compute the interventional effect from observational data. In this particular case, the set of variables containing *x*_1_ and *u*_1_ block all backdoor paths leading from *x*_2_ to *y*. Thus, the interventional distribution is given by

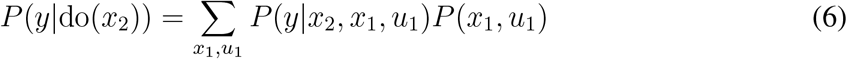

where the do(*x*_2_) function indicates that the value of *x*_2_ is set independently of the other variables. From the previous section, we have assumed that *x*_1_ and *u*_1_ are independent and thus *P* (*x*_1_, *u*_1_) = *P* (*x*_1_)*P* (*u*_1_). However, this assumption could be relaxed by introducing models with multiple patient-level random effects with patient-level predictors as a subset of *x*_1_. Regardless, we see that, under assumptions of the graphical model presented here, conditioning a model for the response of *y* to *x*_2_ on observational data will recover an unbiased estimate of the interventional effect of *x*_2_ on *y*. Insofar as these assumptions are biased, the resulting casual estimates may be as well. Calibration against randomized experiments can quantify the degree of bias in the overall model.

### Real world data from a brain cancer registry

While most clinical research is led by physician-scientists, advocacy- and patient-led research initiatives have become more common with examples reported in the literature (*25*). Advocacy-led research can provide a critical boost especially in areas that are understudied. In particular, advocacy-led registries can help establish the natural history of rare diseases and lay the foundation for development of therapeutics (*26, 27*).

One of the earliest examples of an advocacy- or foundation-led research registry is the Musella Foundation for Brain Tumor Research & Information’s Virtual Trial Registry (hereafter referred to as the virtual trial or VT data) (*28*), which was an advocacy-led initiative begun by the foundation as an on-line forum in 1992, and converted to a web site in 1993, hosted at the Foundation’s site: “virtualtrials.org” (formerly “virtualtrials.com”). Predating the widespread availability of patient resources for this population, the site was developed to provide a clearinghouse where patients with primary brain tumors could share details about their diagnosis, treatments, and health status in order to create a repository of information that could assist other patients.

This registry is an exploratory and descriptive cohort of 761 primary brain tumor patients who self-reported details of their disease and treatments, including treatment outcomes in the form of both tumor scan summaries and Karnofsky performance status (KPS), a standard functional measure for primary brain tumors that is used clinically to stratify patient prognosis and determine appropriate management in glioblastoma multiforme (*29*). Patients provided data voluntarily through the VT web site, and many participants copied details from their medical records into the registry web page.

A common challenge faced by registry studies is the completeness and quality of the data collected. While self-report information can be subject to various biases (*30*), the VT study data was subject to an evaluation in which a pathologist and neurosurgeon independently evaluated patient-reported data against medical charts for a randomly selected sample, which demonstrated significant agreement between patient and physician reported data in this registry (*28*).

We defined a cohort for the present analysis that included patients with a primary brain tumor diagnosis, including glioblastoma, (GBM), anaplastic astrocytoma (AA), diffuse intrinsic pontine glioma (DIPG), and oligodendroglioma (both high and low grades) who reported at least one tumor scan summary, and at least one treatment. Patients were treated between 1980 and 2017 with over 200 different therapies, including cytotoxic chemotherapies, radiation therapy, surgery (partial and/or complete resections), anti-angiogenesis agents, immunotherapy agents, and a small number of targeted therapies. The resulting cohort included data for 451 patients.

To define treatment time intervals, we identified tumor scan summaries in which disease progression (or KPS *<* 10 as a proxy for death) was identified to have occurred. Disease progression was self-assessed by the patient, though in many cases the text of the radiologist’s impression was available for review. Many patients (∼ 40%) had more than one treatment time interval identified, as they were treated again after experiencing disease progression. “Survival” time was defined as the time between the first treatment within a treatment time interval and disease progression.

We chose patient-level predictors for the survival timescale, *µ*, to be tumor type, tumor grade, and age at diagnosis. We chose treatment-level predictors for *µ* to be the resectability of the tumor (partial/complete/unable), patient function (in KPS) at the start of treatment, time since diagnosis, and binary indicators for 35 different treatments. Several of these treatments represent categories of treatment (e.g., “temozolomide” is an instance of a “chemotherapy agent”), which were then coded as additional treatments (e.g., “treated with temozolomide” implies “treated with chemotherapy agent”). We normalized treatment names and resolved treatment categories against the National Cancer Institute thesaurus (*31*).

Chemotherapy agents included carmustine (delivered both intravenous and implanted on the tumor), carboplatin, cisplatin, etoposide, hydroxyurea, irinotecan, lomustine (CCNU), procarbazine, temozolomide (TMZ), vincristine. Anti-angiogenesis agents included bevacizumab, genistein, and thalidomide. Immunotherapy agents included Poly ICLC and several varieties of vaccines. Targeted therapies included erlotinib, imatinib, and tamoxifen. Forms of radiation therapy (RT) included radiosurgery (both fractionated and single dose), intensity-modulated radiation therapy (IMRT), proton beam radiation therapy (PBRT), stereotactic radiosurgery (SRS), conformal radiation therapy, and internal radiation therapy (brachytherapy). Other, unclassified treatments that were considered included celecoxib, dexamethasone, polysaccharide K (PSK), Optune, and valproic acid.

For patient/treatment-level predictors for the variability in survival times, *σ*, we restricted the predictors to be the patient biomarkers, namely tumor type/grade, age, resectability, KPS, and time since diagnosis. For the treatment-level predictors for *µ*, we additionally included pairwise interaction terms between all predictors that had any overlap. To mitigate the effect of multicollinearity, we dropped interaction terms that were entirely or mostly predictive of one of the original terms (e.g., we would have dropped the `radiotherapy+dexamethasone` interaction term if every instance of dexamethasone treatment co-occurred with radiation therapy).

### Fitting the model

We implemented our Bayesian multilevel survival model in the probabilistic programming language, *Stan* (*32*), and fit it to the VT data, using the No-U-Turn Markov Chain Monte Carlo (MCMC) sampler (*33*) to draw samples from the posterior probability distribution. Figure 3 shows a selected number of resulting effect size estimates. Several known clinical insights can be found in these inferred values. Comparing the effect of different conditions, we see that glioblastoma (GBM) is associated with a factor of ∼ 0.5 decrease in expected survival, compared to anaplastic astrocytoma (AA, ∼ 1) and oligodendroglioma (∼ 1.5). Treatment that occur within three months of diagnosis (indicated as “newly diagnosed”) are associated with a factor of *>* 2 longer time to progression, compared to treatment that occurs later, possibly when the patient has experienced recurrence or previous treatment failure.

**Figure 3:**
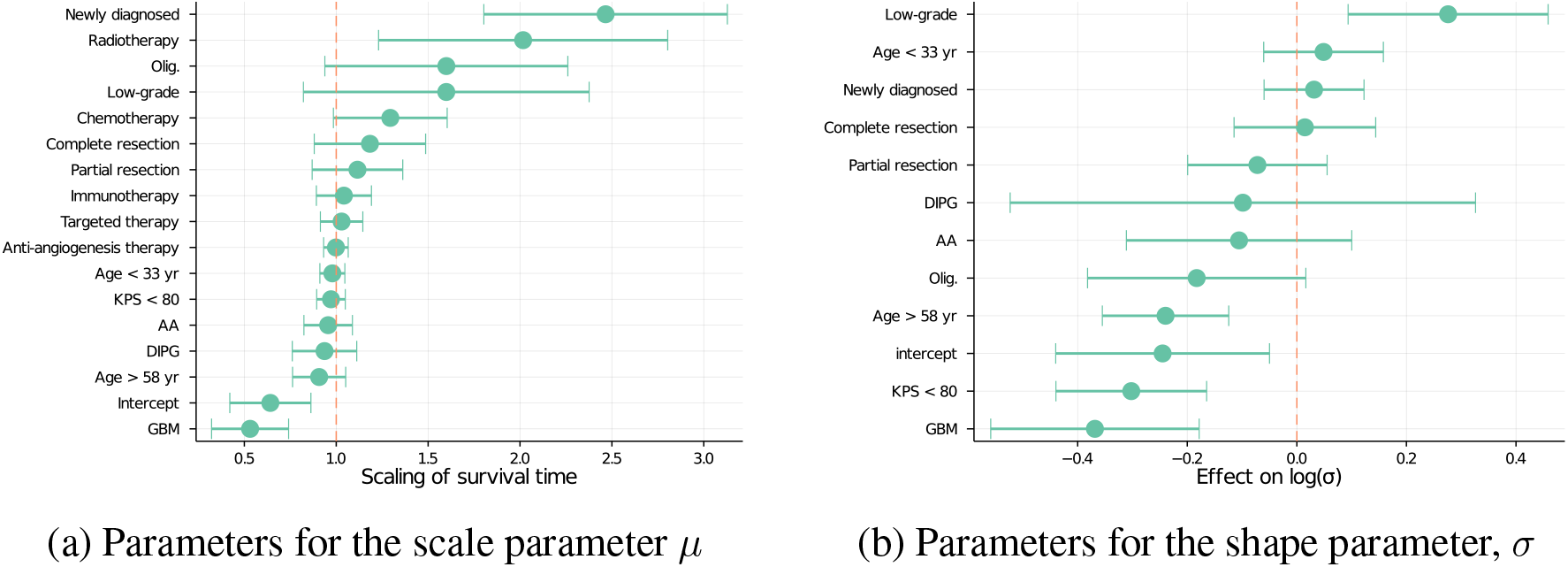
Posterior mean and standard deviations for effect size parameters in the model as fitted to the Musella VT data. The left panel shows a subset of the effect sizes for *µ*, transformed with an exponential so as to represent the effect on the acceleration factor of the survival time. The right panel shows the effect sizes for log *σ*, which quantifies the degree of variance about expected survival times. We see that often (but not always), patient features that predict shorter survival times also predict lower variance in these worse outcomes.

### Causal validation

The model presented in the prior sections contains various causal assumptions, summarized in the graphical diagram in Figure 2. Verifying that all of these assumptions hold is infeasible in practice, and so to assess the degree of causal bias in the observational model, we compared our model’s estimated treatment effects with those found in RCTs. The model development and analyses were completed before any RCTs were considered for comparison, and the results of these RCTs were not known at the time of model development. We queried ClinicalTrials.gov for RCTs matching the VT patient population (at least one of glioblastoma, anaplastic astrocytoma, diffuse intrinsic pontine glioma, or oligodendroglioma), and with at least two treatment arms represented in the registry. These queries resulted in a list of seven comparison RCTs (described in Table 1) (*34–40*). In total, there were over 3200 participants across these seven clinical trials.

**Table 1:** List of randomized controlled trials for comparison with the results of the virtual trial.

| Trial ID | Publication | $N_{\text{pt}}$ | Condition | Arm A | Arm B |
| --- | --- | --- | --- | --- | --- |
| NCT01149109 | (40) | 141 | Newly diagnosed glioblastoma | temozolomide, lomustine | temozolomide |
| NCT01290939 | (39) | 437 | Progressive glioblastoma | lomustine, bevacizumab | lomustine |
| NCT00182819 | (38) | 477 | Low-grade glioma | temozolomide | radiotherapy |
| NCT00884741 | (37) | 978 | Newly diagnosed glioblastoma | temozolomide, radiotherapy, bevacizumab | temozolomide, radiotherapy |
| NCT00943826 | (36) | 921 | Newly diagnosed glioblastoma | temozolomide, radiotherapy, bevacizumab | temozolomide, radiotherapy |
| NCT00052455 | (35) | 447 | Recurrent high-grade glioma | procarbazine, lomustine, vincristine | temozolomide |
| NCT00002569 | (34) | 289 | Newly diagnosed high-grade oligo-dendroglioma or oligoastrocytoma | procarbazine, lomustine, vincristine, radiotherapy | radiotherapy |

For each RCT, we used the model as fitted with the VT data to make predictions for the median time-to-progression experienced by participants within each treatment arm. It should be noted that the predicted endpoint will, by construction, differ from an endpoint of PFS from a randomized trial, as the definition of a starting time is different between the VT and an RCT. We marginalized the predicted survival times over both the posterior probability distribution and a uniformly random distribution of predictors, with particular entries fixed to be consistent with the inclusion/exclusion criteria of the trial and the specific treatment arm. Figure 4 shows the resulting ratios in median survival between treatment arms, both observed PFS endpoints from the RCT and the VT prediction. For all but one of the seven comparison trials (namely the PCV vs TMZ trial described by (*35*)), the VT model reproduces the direction of the observed ratio of PFS between treatment arms, and five of the seven comparisons show that the mean VT estimated log PFS ratio lies within the 95% confidence interval of the corresponding trials.

**Figure 4:**
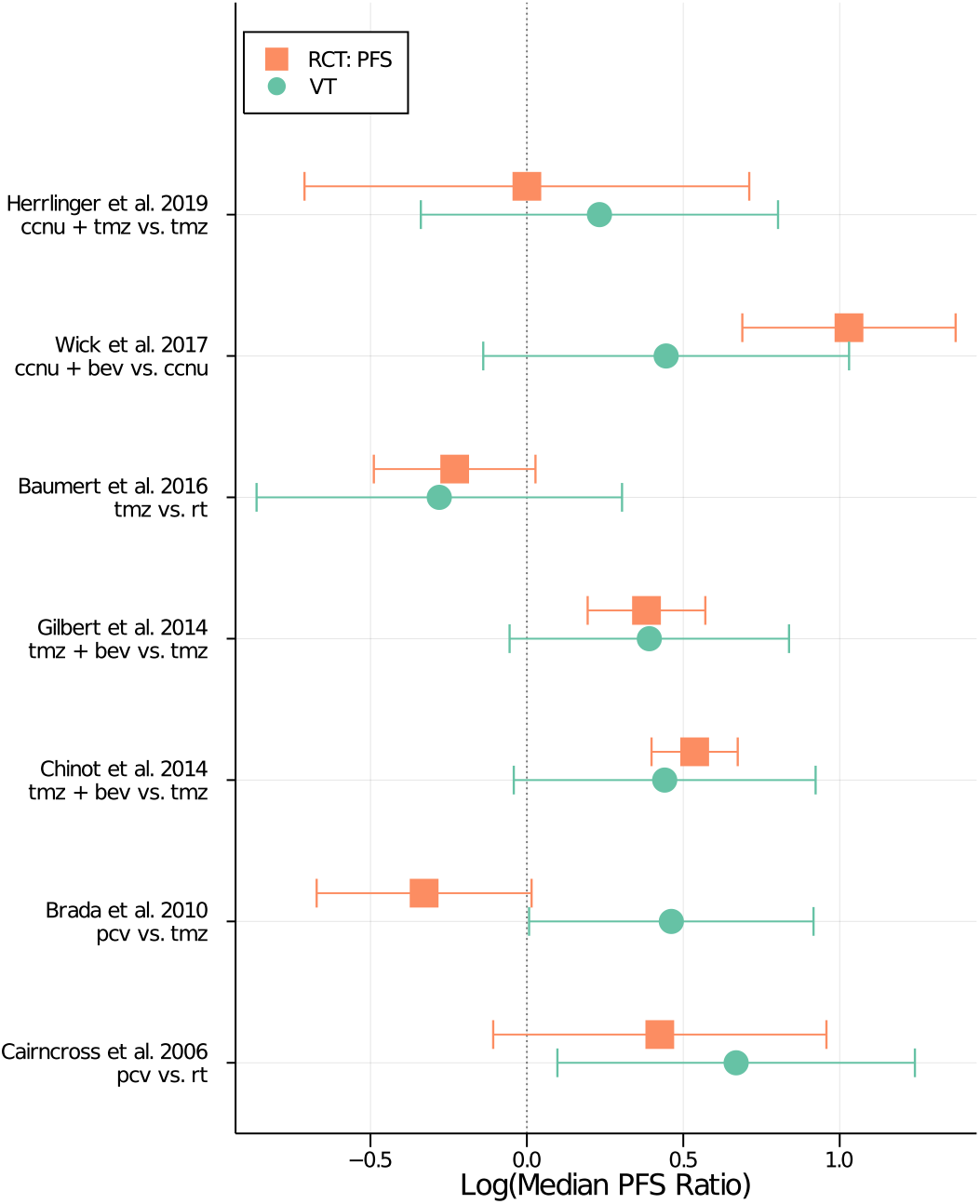
Comparison of simulations from the virtual trial (VT) model to results from a set of randomized controlled trials (RCTs). The *x*-axis shows the logarithm of the ratio of median progression-free survival (PFS) times between treatment arms. The *y*-axis shows different RCTs, with progression-free survival (PFS) results from the trial in orange squares, and the simulated result from the VT model in green circles. Treatments have been abbreviated as follows: lomustine is “ccnu”, temozolomide is “tmz”, bevacizumab is “bev”, radiation therapy is “rt”, and procarbazine-lomustine-vincristine is “pcv”. The 95% confidence interval for the log ratio of RCT survival times was approximated using linear error propagation, while the 95% credible interval for the log ratio of the VT simulations was calculated by marginalizing over the posterior probability distribution and random realizations of the patient sample.

### Resolving the PCV vs TMZ discrepancy

The most notable difference between the model-predicted treatment effects and those from RCTs was found for the PCV vs TMZ comparison. One key difference between the patient population in this study and our registry was that the PCV vs TMZ study was restricted to chemotherapy-naive recurrent glioma patients. In contrast, patients in our registry who had recurrent high grade glioma generally received upfront chemotherapy, following standard of care. Brada and colleagues (*35*) concluded that there was no difference between PCV and TMZ for high grade glioma on the PFS and OS endpoints, but with the simpler dosing schedule and favorable tolerability, use of TMZ eclipsed PCV for this population in the years that followed. However, PCV has many advocates and the literature has evolved with several new biomarkers that were not available in the Brada trial or in our registry. In a 2018 review, Hafazalla et al. (*41*) summarized the situation as follows, “the data suggest that for patients harboring a tumor with an unfavorable natural history, such as those with intact 1p/19q and wild-type IDH1, TMZ and RT may be the best option. Conversely, the data suggest that patients with biologically favorable LGG are likely to derive the most significant benefit from RT and adjuvant PCV. A prospective trial directly comparing PCV and TMZ in patients with high-risk low-grade glioma is needed.” Indeed, several large RCTs are underway that are expected to produce results in late 2025 (see, e.g. NCT00887146). Our results may thus be interpreted as a reason to continue investigating the ideal population or sub-population.

### Predicted outcomes for new patients

In addition to summarizing population-level treatment effects, we can use the model to estimate survival distributions for individual patients, conditioned on their features and proposed treatment. These estimated survival distributions can then be used to help inform patient treatment decisions. To demonstrated this task, we generated synthetic patient cases using various patient features from the Musella VT dataset. We then proposed a set of plausible treatments (including single agents and combinations). For each patient and each proposed treatment, we sampled first from the posterior probability distribution over the model parameters and then from the survival likelihood as conditioned on those model parameters. By repeating this sampling procedure for a large number of iterations, we built up a distribution of individual survival time predictions that can be compared to other patients and other treatments. Figure 5 shows the mean and standard deviation of these predictions for three sample patients across a set of proposed treatments. While there is a large overlap in the credible intervals for treatment effects, the order of the mean treatment effects differ from patient to patient. For instance, for a low-grade oligodendroglioma patient, radiation is preferred over the combination of bevacizumab and lomustine, while this order is reversed for a glioblastoma patient with low performance status.

**Figure 5:**
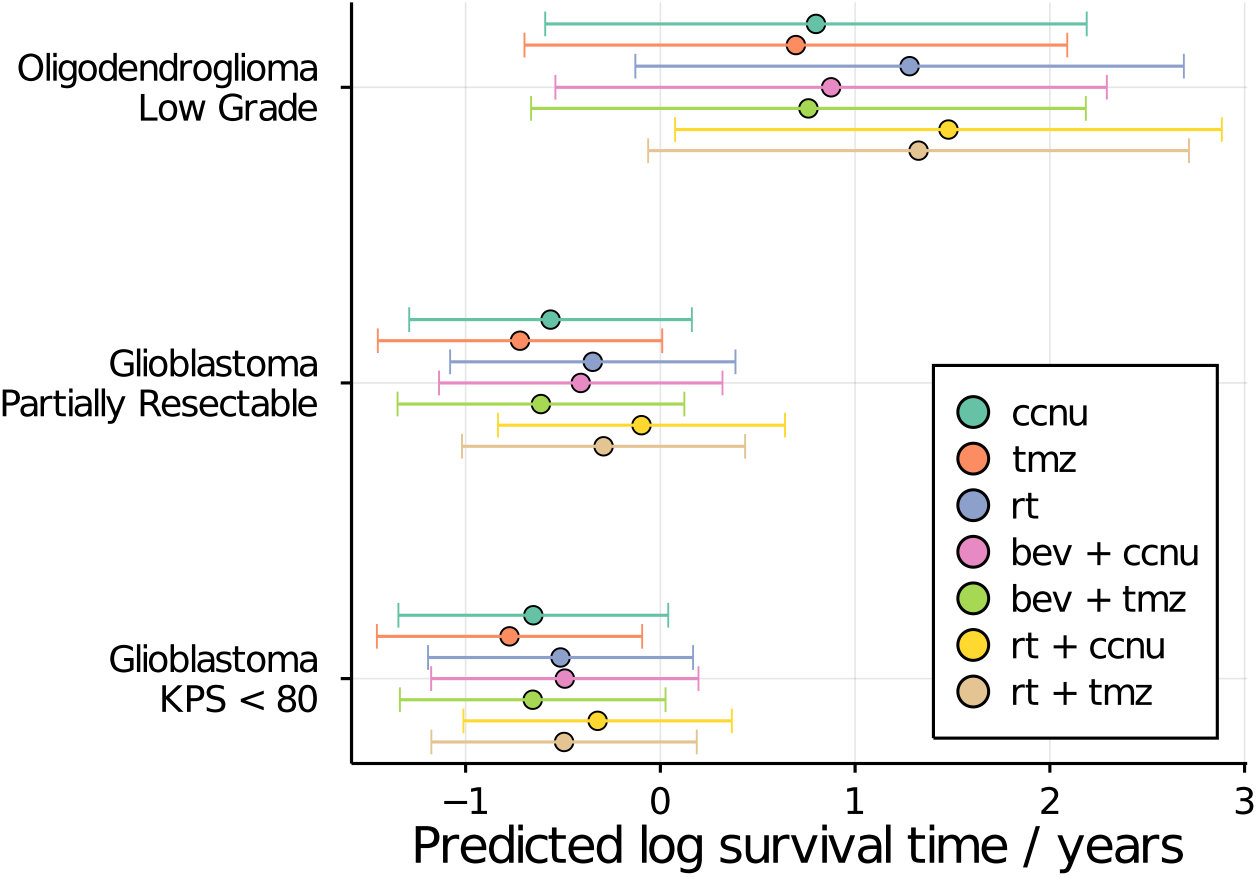
Posterior predictive survival distributions conditions on various patient features. Each row shows a different synthetic patient case with the features listed on the *y*-axis. The effect of different treatments on survival for each patient are shown as the various colored errorbars. While the credible intervals largely overlap with one another, the order of the mean treatment effects across different treatment combinations vary from patient to patient.

## Discussion

Randomized clinical trials with narrow inclusion criteria are necessary to satisfy the hypothesis testing requirement of an independent and identically distributed population; but as the inclusion criteria become more narrow, finding large enough populations to make meaningful inferences becomes more difficult. This limitation both slows the accrual rate of clinical trials and excludes many cancer patients from trials, thus slowing the pace of knowledge generation. Slow enrollment leads to higher trial costs. This paradigm had resulted in a greater emphasis on measures of treatment effect rather than clinically-actionable information on patient features that moderate or mediate treatment effects. A consequence of this is that large numbers of cancer patients lose out on the opportunity and potential benefits of clinical research (*42*).

The speed and cost of oncology drug development could be reduced by strategies that allow for more inclusive approaches to patient enrollment and inference across a larger and more diverse patient population. This could be accomplished by a greater reliance on real-world data, which would reduce the burden that clinical research places on busy clinicians–another factor limiting enrollment into clinical trials.

The model we present provides a tool for extracting clinically meaningful information from heterogeneous real-world data. For example, in glioma, where there is a diversity of clinical practice, the model can help identify the ideal therapy for an individual considering a variety of biomarkers. Indeed our motivation was to explore whether a single machine-learning model could recover the effects of known clinical biomarkers and treatment effects from a medium-sized, non-randomized sample. We believe that the data presented shows the method promising for use is precision oncology, where the number of biomarkers, treatments, and treatment/biomarker interactions may lead to a decades-long search for the optimal regimen for specific subtypes. This is important for identifying the optimal use of approved therapies efficiently from registries of clinical practice that are less costly than large, global RCTs. For example NCT00887146, is expected to take 16 years and require 271 global sites to answer the PCV versus TMZ use question.

A natural concern for using models trained on observational data to drive clinical decision making is the potential for biases in the estimated treatment effects. Indeed, a recent comparison of observational studies with oncology RCTs by Soni et al. (*43*) found agreement that was only marginally better than the agreement from random chance. However, this analysis was restricted to studies with overall survival endpoints, which the authors noted are less reliable than intermediate endpoints such as PFS or time to treatment failure. Furthermore, larger meta-analyses of comparisons between observational studies and RCTs across a diverse range of clinical domains have found little difference, on average between the two experiment types (*44*). In the specific setting of oncology, Petito et al. (*45*) applied a targeted trial approach to observational data from SEER/Medicare, one of the data sources used by Soni et al. The authors’ SEER/Medicare-derived estimates were in good agreement with the two targeted RCTs. However, as Petito and colleagues noted, the agreement was dependent on their proper accounting for censoring and confounding (via inverse probability weighting). Clearly there are challenges in the design of non-randomized experiments, but we believe that our results realize the efficiency benefits of observational studies, without substantial bias with respect to RCTs.

## Conclusion

We have demonstrated that it is possible to learn features and feature-by-treatment interactions from heterogeneous real-world cancer data, and that effect size measures from this approach generally recapitulate treatment effects sizes from randomized clinical trials, without requiring a carefully selected population, and with more information learned from a smaller overall dataset. This model also facilitates individual-level forecasts that incorporate large clinical feature sets and provides easily interpretable measurements of parameters, both requirements for clinically useful precision oncology tools. This is an important step towards maximizing information gain from clinical medicine and better integrating clinical research into the practice of medicine.

## Data Availability

The Foundation is seeking an IRB exemption to release the de-identified disaggregated data used in the present research. If The Foundation obtains that exemption, it will make the de-identified disaggregated data publicly available. Until such time as the de-identified disaggregated data is publicly released, it can be obtained through case-by-case IRB approval. Individuals wishing to obtain the de-identified disaggregated data should contact either the corresponding author (Asher Wasserman), or the Musella Foundation.

https://virtualtrials.org/musella.cfm

## Status of Data and Availability

The dataset used in this work was assembled by The Musella Foundation for Brain Tumor Research (virtualtrials.org). Patients enrolling in The Foundation’s observational Brain Tumor Virtual Trial Study, or their advocates, voluntarily submitted information about themselves and their treatments and test results to The Foundation. Patients were specifically consented for the collection of this data, and agreed that the data could be released in aggregate analyses, such as the present one. This consent was approved by The Musella Foundation’s oversight board. The Foundation is seeking an IRB exemption to release the de-identified disaggregated data used in the present research. If The Foundation obtains that exemption, it will make the de-identified disaggregated data publicly available. Until such time as the de-identified dis-aggregated data is publicly released, it can be obtained through case-by-case IRB approval. Individuals wishing to obtain the de-identified disaggregated data should contact either the corresponding author (Asher Wasserman), or the Musella Foundation.

## Acknowledgments and Conflicts

We thank Glenn Kramer, Kristian Thorlund, and Jameson Quinn for their comments and feedback. In addition, we thank the reviewers and editors for thoughtful comments that improved the quality of this paper. This work was supported by xCures, Inc. Portions of the material described here are included in a patent application describing a computer-implemented system.

## References

1. D. Chakravarty, et al., JCO Precision Oncology pp. 1–16 (2017).

2. J. P. Jansen, et al., Value in health : the journal of the International Society for Pharmacoeconomics and Outcomes Research 14, 417 (2011).

3. J. Shrager, M. Shapiro, W. Hoos, The Journal of Law, Medicine & Ethics 47, 362 (2019).

4. S. J. Niranjan, et al., Cancer 126, 1958 (2020).

5. J. M. Unger, R. Vaidya, D. L. Hershman, L. M. Minasian, M. E. Fleury, JNCI: Journal of the National Cancer Institute 111, 245 (2019).

6. T. Hampton, JAMA 297, 683 (2007).

7. R. Rahman, et al., Clinical Cancer Research 25, 6339 (2019).

8. Y. Li, M. Liang, L. Mao, S. Wang, arXiv:1912.09664 [stat] (2019).

9. L. Lapointe-Shaw, et al., BMC Medical Research Methodology 18, 118 (2018).

10. C. A. Bellera, et al., BMC Medical Research Methodology 10, 20 (2010).

11. N. Simon, J. Friedman, T. Hastie, R. Tibshirani, Journal of statistical software 39, 1 (2011).

12. Z. Zhang, J. Reinikainen, K. A. Adeleke, M. E. Pieterse, C. G. M. Groothuis-Oudshoorn, Annals of Translational Medicine 6 (2018).

13. J.-A. W. Chapman, et al., Breast Cancer Research and Treatment 22, 263 (1992).

14. Z. Iraji, T. Jafari Koshki, R. Dolatkhah, M. Asghari Jafarabadi, Journal of Research in Medical Sciences : The Official Journal of Isfahan University of Medical Sciences 25 (2020).

15. N. Keiding, P. K. Andersen, J. P. Klein, Statistics in Medicine 16, 215 (1997).

16. M. J. Crowther, P. Royston, M. Clements, 2006.06807 [stat] (2020). Tex.ids= crowther2020a.

17. L. J. Wei, Statistics in Medicine 11, 1871 (1992). Tex.ids= wei1992a publisher: John Wiley & Sons, Ltd.

18. A. Sashegyi, D. Ferry, The Oncologist 22, 484 (2017).

19. J. K. Kruschke, T. M. Liddell, Psychonomic Bulletin & Review 25, 178 (2018).

20. D. G. Kleinbaum, M. Klein, Survival analysis: a self-learning text, Statistics for biology and health (Springer, New York, 2012), third edn.

21. A. Gelman, Bayesian Analysis 1, 515 (2006).

22. A. Gelman, J. Hill, Data analysis using regression and multilevel/hierarchical models, Analytical methods for social research (Cambridge University Press, Cambridge ; New York, 2007).

23. J. Piironen, A. Vehtari, 1610.05559 [stat] (2017).

24. J. Pearl, Causality (Cambridge University Press, 2009), second edn. Google-Books-ID: f4nuexsNVZIC.

25. S. F. Terry, P. F. Terry, K. A. Rauen, J. Uitto, L. G. Bercovitch, Nature Reviews Genetics 8, 157 (2007).

26. D. C. Landy, et al., Genetics in Medicine 14, 223 (2012).

27. J.-M. Quach, R. Campbell, J. Walsh, Human Gene Therapy 26, 783 (2015).

28. R. Hayes, et al., 90th meeting of the Am. Assoc. for Cancer Research (AACR, Philadelphia, PA, 1999), pp. 39–40.

29. L. B. Chambless, et al., Journal of Neuro-Oncology 121, 359 (2015).

30. R. Gliklich, M. Leavy, N. Dreyer, Registries for Evaluating Patient Outcomes: A User’s Guide (Agency for Healthcare Research and Quality, 2020), fourth edn.

31. J. Golbeck, et al., Journal of Web Semantics 1 (2003).

32. B. Carpenter, et al., Journal of Statistical Software 76, 1 (2017).

33. M. D. Hoffman, A. Gelman, Journal of Machine Learning Research 15, 31 (2014).

34. G. Cairncross, et al., Journal of Clinical Oncology 24, 2707 (2006).

35. M. Brada, et al., Journal of Clinical Oncology 28, 4601 (2010).

36. O. L. Chinot, et al., New England Journal of Medicine 370, 709 (2014).

37. M. R. Gilbert, et al., n engl j med p. 10 (2014).

38. B. G. Baumert, et al., The Lancet. Oncology 17, 1521 (2016).

39. W. Wick, et al., n engl j med p. 10 (2017).

40. U. Herrlinger, et al., The Lancet 393, 678 (2019).

41. K. Hafazalla, A. Sahgal, B. Jaja, J. R. Perry, S. Das, Oncotarget 9, 33623 (2018).

42. L. Rodriguez, P. Lowy, Protocol Complexity and Patient Enrollment Intensify Challenges in Oncology Clinical Trials, According to Tufts Center for the Study of Drug Development (2021).

43. P. D. Soni, et al., Journal of Clinical Oncology: Official Journal of the American Society of Clinical Oncology 37, 1209 (2019).

44. A. Anglemyer, H. T. Horvath, L. Bero, Cochrane Database of Systematic Reviews (2014). Publisher: John Wiley & Sons, Ltd.

45. L. C. Petito, et al., JAMA network open 3, e200452 (2020).

